# Projected impact on blood pressure, chronic kidney disease burden and healthcare costs of achieving the Australian sodium reduction targets: a modelling study

**DOI:** 10.1101/2022.03.18.22272639

**Authors:** Leopold N. Aminde, Mary Wanjau, Linda J. Cobiac, J. Lennert Veerman

**Author notes:** **Correspondence:** Dr. Leopold N. Aminde, School of Medicine and Dentistry, Griffith University, Ian O’Connor Building (G40), Gold Coast campus, 1 Parklands Drive, QLD 4215, Australia. or.

## Abstract

**Background:** Excess sodium intake increases blood pressure and risk of vascular complications. Most studies have focused on heart disease and stroke, but the impact on chronic kidney disease (CKD) is less well-characterised. The aim of this study was to estimate the impact of sodium intake on CKD burden in Australia.

**Methods:** A dynamic epidemiological model was developed to estimate the potential impact on CKD if the Australian Suggested Dietary Target (SDT) and the National Preventive Health Strategy 2021-2030 (NPHS) sodium target were achieved. Outcomes were estimated between 2019 (base year) and 2030, and lifetime.

**Findings:** Achieving the SDT and NPHS target could lower overall mean systolic blood pressure by 2.1 mmHg and 1.7 mmHg respectively. Compared to current sodium intake levels, attaining the SDT could prevent 59,220 (95% UI, 53,140 – 65,500) incident CKD cases and 568 (95% UI, 479 – 652) CKD deaths by 2030. Over the lifetime, this is projected to generate 199,488 (95% UI, 180,910 – 218,340) HALYs and AU$ 644 million in CKD health expenditure savings. Similarly, if achieved, the NPHS is estimated to prevent 49,890 (95% UI, 44,377 – 55,569) incident CKD cases and 511 (95% UI, 426 – 590) CKD deaths by 2030. Over the lifetime, this could produce 170,425 (95% UI, 155,017 – 186,559) HALYs and AU$ 514 million in CKD health expenditure savings.

**Conclusion:** Achieving the national sodium reduction targets could deliver substantial health and economic benefits for people with CKD in Australia. Robust government action is required in order to achieve the projected outcomes of these policy targets.

## INTRODUCTION

Excess sodium intake is the leading dietary risk for death worldwide. A recent global analysis showed that diets high in sodium were responsible for more than 3 million deaths and over 70 million disability-adjusted life years (DALYs) (1). One major way through which excess sodium intake affects health is by raising blood pressure (BP). The INTERSALT study that included over 10,000 adults from 32 countries was the earliest large-scale epidemiological study that demonstrated that high urinary sodium excretion (a proxy for intake) and a high sodium-potassium ratio assessed via 24-hour urine were associated with increases in BP (2). Meta-analyses of randomized trials have confirmed a robust linear and dose-response relationship between sodium intake and BP. This causal effect is stronger in those with hypertension, black ethnic populations, and greater for systolic BP than diastolic BP (3, 4).

In Australia, Huggins and colleagues were the first to demonstrate this link between sodium and BP. Using data from the Melbourne Collaborative Cohort Study (MCCS), they found that after adjusting for age, body mass index, country of birth and anti-hypertensive medication use, a 100 mmol/day increase in sodium was associated with a 2.3 mmHg increase in systolic BP (5). Furthermore, the recent National Health Survey 2017-18 revealed that 21% of the cases of hypertension in Australia were due to diets high in sodium (6).

Lowering sodium intake in the population reduces average BP and slows the process of atherosclerosis, and the advancement of kidney injury to End Stage Kidney Disease (8, 9). A recent Cochrane meta-analysis of randomized trials in people with CKD demonstrated that a reduction in salt intake by 4.2g (1,690mg sodium) per day lowered systolic and diastolic BP by 6.91 (4.99 – 8.82) mmHg and 3.91 (3.02 – 4.80) mmHg, respectively. In addition, for people with early stages of CKD, this salt reduction led to a 36% (26 – 44) reduction in albuminuria, a potent marker of poor kidney function (10). Therefore, decreasing average population sodium consumption could play a significant role in reducing the morbidity and mortality associated with CKD in Australia.

In 2019, an estimated 62% of the CKD deaths and 56% of CKD-related DALYs lost in Australia were due to hypertension (11). Despite compelling evidence on the relationship between dietary sodium, BP and CKD, few studies have assessed the long-term impact of sodium reduction on CKD (12). Most modelling studies globally (13-17), and in Australia (18, 19) have either focused on the impact on heart disease and stroke or did not explore the longer-term impacts (20). This implies the currently reported health impacts of sodium reduction strategies in Australia are underestimated. The aim of this study was to complement this knowledge gap by developing a model to quantify the impacts of dietary sodium reduction on the future burden of CKD, and related health expenditure in Australia.

## METHODS

### Specification of modelled salt targets

Two main sodium policy targets are modelled in this study: the Australian Suggested Dietary Target (SDT) and the National Preventive Health Strategy (NPHS) 2021-2030 target (21, 22). The National Health and Medical Research Council developed nutrient reference values (NRV) for a range of nutrients including sodium in 2006 and updated these in 2017 to reflect best contemporary evidence. As a result, the SDT, defined as *a daily average intake from food and beverages for certain nutrients that may help in prevention of chronic disease* was established for sodium as 2,000mg (5g of salt) per day (21). The recent Australian National Preventive Health Strategy 2021-2030 specifies a long-term approach to prevention and identifies key focus areas for the next 10 years. With respect to healthy diets, a key target is to reduce sodium intakes by an average of 30% by 2030 (22). In this analysis, we compared the impact on CKD of current population sodium intake with a counterfactual (attaining the policy targets), assuming a gradual linear reduction over ten years towards the respective targets.

### Sodium intake and blood pressure data

Data on sodium intake was obtained from a meta-analysis of 31 published studies and 1 unpublished dataset of surveys conducted across Australia between 1989 to 2015. After adjusting for non-urinary excretion, the overall weighted average daily intake was ∼3700mg of sodium (9.6g of salt) based on 24-hour urine collections (23). This meta-analysis included results from the 2011-12 National Nutrition and Physical Activity Survey (NNPAS) (24), a nationally representative survey of over 12,000 adult Australians and estimated sodium intake using 24-hour dietary recall. We used the weighted average 24-hour urine sodium estimates from the meta-analysis in our modelling and derived age- and sex-specific trends in sodium intake from the NNPAS to adjust the overall estimates accordingly. This was done to account for the age-related differences in baseline sodium intake as we modelled the relationship with blood pressure by multiple age-cohorts.

Data on blood pressure (BP) was from the Australian 2017-18 National Health Survey (25). This nationally representative survey used a random multistage area sampling of private dwellings, with a final sample of 16,384 dwellings and 21,315 participants. It was designed to collect data on chronic health conditions and risk factors like BP, obesity, smoking, alcohol consumption and physical activity. Consenting adults (18 years and above) had 3 BP measurements which were averaged. Estimates of the weighted mean measured BP by age-groups and sex were used for our analysis. *See supplementary file for all input data*.

#### Model framework

We used the proportional multistate lifetable (PMSLT), a dynamic epidemiological model suited for simulating long-term population impacts of interventions for chronic disease prevention. This analysis took a three-step approach: first, the effect of a changes in sodium on BP; second, the effect of changes in blood pressure on incident CKD; and finally, the effect of changes in CKD morbidity and mortality on health-adjusted life years (HALY) and health expenditure.

### Sodium, blood pressure and epidemiological modelling

The change in systolic BP from a change in sodium consumption was based on a Cochrane meta-analysis of randomized trials of interventions conducted over at least a month, as these are considered relevant from a public health perspective (26). This meta-analysis demonstrated that reduction by 4.4g salt (∼1,720mg sodium) led to a 5.39 (95% CI 4.15 to 6.62) mmHg and 2.42 (1.29 to 3.56) mmHg drop in systolic BP for people with and without hypertension respectively. We accounted for this differential impact of changes in sodium for people with mean systolic BP above and below 140mmHg.

Data on the relative risks of CKD due to changes in systolic BP and baseline data on the epidemiology of CKD in Australia were obtained from the 2019 Global Burden of Disease (GBD) study (11). DISMOD-II epidemiological software was used to derive CKD case fatality rates. This analytical software uses a set of differential equations that exploit the causal relation in a typical disease process to estimate absent epidemiological parameters while maintaining stability in the overall disease epidemiology (27). We calculated the impact of the resulting change in SBP on CKD using the potential impact fraction (PIF) (28). By integrating the risk function with the exposure (BP) distribution for each 5-year age cohort by sex, with and without the intervention (sodium reduction), the PIF quantified the proportional change in the incidence of CKD. The ‘distribution shift’ method of the PIF calculation was used in this analysis (*see supplementary file for details*).

### Multistate lifetable modelling

We modelled the different CKD causes (CKD due to hypertension, CKD due to diabetes mellitus, CKD due to glomerulonephritis and CKD due to unknown causes) as implemented in the GBD (29). Each condition was modelled as a separate Markov sub-model in the PMSLT, in which proportions move between four Markov health states: Alive without CKD, Alive with CKD, Dead from CKD and Dead from other causes, in 1-year cycles. These transitions are informed by incidence and case fatality hazards, with death an absorbing state. Given the progressive nature of CKD, we assumed zero remission. The disease Markov models were linked to the lifetable. The PIF described above calculates new (post-intervention) incidence rates of CKD which leads to changes in prevalence, with mortality being a function of prevalence. Mortality propagates into the lifetable which recalculates all-cause mortality and life years. Life years are adjusted for loss of quality life due to CKD for each age and sex to obtain HALYs. One HALY could thus be defined as a year of life in perfect health *(see supplementary file)*. Five-year age-group and sex-specific simulations are conducted for the remaining lifetime or until people reach 100 years, simultaneously for the reference and intervention populations. The difference in health outcomes (incidence, mortality and HALYs) between the two populations quantifies the impact of the policy targets.

### Healthcare costs

Cost data for CKD were obtained from the Australian Institute of Health and Welfare estimates for health expenditure 2018-2019 (30). We divided total annual health spending for CKD by the corresponding prevalent numbers to obtain the annual per capita costs for each sex and five-year age group. The resulting estimates were multiplied by the change in prevalent cases of CKD due the policy targets to estimate the impacts on health expenditure. Our model base year was 2019 so we did not need inflation adjustments. Costs projections beyond the base year were discounted (31).

### Scenario and uncertainty analysis

Our main analysis included: i) reducing current sodium intake to an average of 2,000mg of sodium or 5g of salt (Australian SDT) and ii) Reducing average sodium intake by 30% (NPHS, 2021-2030). In addition, we conducted the following scenario analyses: i) a 20% relative reduction in sodium intake, ii) a 15% relative reduction in sodium intake (similar to that achieved in the UK salt reduction program), iii) a reduction in salt intake by 1g, and iv) a reduction in salt intake by 2g. Our main analysis presents undiscounted HALYs while costs are discounted at 3%. We conducted sensitivity analyses with HALYs discounted at 3% and costs discounted at 5% (31).

We conducted probabilistic sensitivity analysis to account for the impact of uncertainty in model input parameters on outcomes. Plausible statistical distributions for parameters with known uncertainty were specified: sodium intake (normal), blood pressure (normal), sodium-BP relationship (normal), relative risks (lognormal), costs (gamma), from which random draws were made using Monte Carlo simulations implemented with Ersatz version 1.35 (32). We ran 2000 simulations and report the 95% uncertainty intervals (2.5^th^ and 97.5^th^ percentiles).

## RESULTS

### Impact of sodium reduction on mean blood pressure

To achieve the Australian SDT by 2030, our model estimates that a modest linear reduction from current sodium levels, that is, on average 0.46g of salt (∼180mg sodium) annually for 11 years could lower mean systolic BP by 2.93 mmHg in men and 1.3 mmHg in women. Secondly, achieving the NPHS target by 2030 would require an average annual decline of 0.29 g of salt (110mg sodium). This could reduce mean systolic BP in 2030 by 1.91 mmHg and 1.42 mmHg for men and women respectively. Table 1 depicts systolic BP distributions with and without the intervention.

**Table 1:** Estimated changes in systolic BP following sodium reduction.

| Age | Men |  |  | Women |  |  |
| --- | --- | --- | --- | --- | --- | --- |
|  | Baseline | Post - SDT | Post - NPHS | Baseline | Post - SDT | Post - NPHS |
| 25 – 34 years | 120.3 (13.5) | 116.6 (12.7) | 118.4 (13.1) | 108.5 (15.8) | 106.7 (15.3) | 107.1 (15.4) |
| 35 – 44 years | 121.3 (11.6) | 118.1 (10.9) | 119.5 (11.2) | 112.5 (15.7) | 111.0 (15.4) | 111.2 (15.4) |
| 45 – 54 years | 126.5 (17.9) | 123.3 (17.1) | 124.7 (17.5) | 119.6 (17.0) | 118.1 (16.6) | 118.3 (16.7) |
| 55 – 64 years | 132.4 (14.7) | 129.9 (14.1) | 130.8 (14.3) | 126.8 (20.6) | 125.6 (20.4) | 125.6 (20.3) |
| 65 – 74 years | 134.9 (21.3) | 132.5 (20.7) | 133.4 (20.9) | 133.6 (17.7) | 132.5 (17.5) | 132.4 (17.4) |
| 75 – 84 years | 136.5 (20.9) | 134.7 (20.5) | 135.1 (20.6) | 137.9 (17.0) | 137.2 (16.7) | 136.8 (16.8) |
| 85 and above | 140.2 (33.3) | 136.2 (32.8) | 137.2 (32.9) | 140.8 (28.0) | 139.2 (27.8) | 138.5 (27.7) |

*Numbers are mean (standard deviation); SDT, Suggested Dietary Target; NPHS, National Preventive Health Strategy 2021-2030 target.*

### Estimated changes in CKD incidence by 2030 and lifetime

Between 2019 and 2030, attaining the SDT could prevent over 59,220 (95% uncertainty intervals [UI]: 53,140 to 65,500) new cases of CKD overall. In the same period, achieving the NPHS target could prevent 49,890 (95%UI: 44,370 to 55,570) new cases of CKD. If the sodium targets were sustained for the remaining lifetime, this would result in 386,800 (95%UI: 347,450 to 427,680) and 348,325 (312,980 to 386,620) incident cases of CKD avoided for the SDT and NPHS target respectively. For both targets and over 11-year and lifetime horizons, most of the avoided new cases were CKD of undetermined causes (53%) followed by CKD due to hypertension (23%). Furthermore, relative reductions in cumulative incident CKD over the lifetime were bigger in men compared to women, that is, 8.3% vs. 3.2% for SDT and 6.1% vs. 3.8% for NPHS. See Table 2, Figure 1 and supplementary file Figures 2 – 5.

**Table 2:** Estimated reductions in cumulative number of incident cases of CKD in Australia between 2019 to 2030, and over the lifetime.

| Reductions between 2019 to 2030 |  |  |  |
| --- | --- | --- | --- |
|  | Male<br>(Mean 95% UIs) | Female<br>(Mean 95% UIs) | Both<br>(Mean 95% UIs) |
| Scenario 1 | 40,518<br>(35,890 - 45,264) | 18,704<br>(16,399 - 21,216) | 59,223<br>(53,140 - 65,503) |
| Scenario 2 | 27,748<br>(24,194 - 31,258) | 22,142<br>(19,494 - 24,975) | 49,890<br>(44,377 - 55,569) |
| Scenario 3 | 18,496<br>(16,182 - 20,817) | 14,774<br>(12,962 - 16,703) | 33,270<br>(29,672 - 36,947) |
| Scenario 4 | 13,846<br>(11,988 - 15,597) | 11,072<br>(9,611 - 12,491) | 24,919<br>(22,141 - 27,724) |
| Scenario 5 | 10,401<br>(8,992 - 11,817) | 11,018<br>(9,625 - 12,578) | 21,419<br>(18,967 - 24,069) |
| Scenario 6 | 20,838<br>(18,037 - 23,603) | 22,050<br>(19,126 - 25,018) | 42,888<br>(37,943 - 47,892) |
| Reductions over the remaining lifetime |  |  |  |
|  | Male<br>(Mean 95% UIs) | Female<br>(Mean 95% UIs) | Both<br>(Mean 95% UIs) |
| Scenario 1 | 266,676<br>(237,912 - 295,250) | 120,133<br>(106,868 - 133,853) | 386,809<br>(347,452 - 427,680) |
| Scenario 2 | 192,140<br>(171,383 - 214,418) | 156,186<br>(138,862 - 174,500) | 348,326<br>(312,986 - 386,619) |
| Scenario 3 | 128,163<br>(113,735 - 142,551) | 103,853<br>(92,544 - 115,616) | 232,016<br>(207,515 - 257,151) |
| Scenario 4 | 96,067<br>(85,977 - 106,793) | 77,820<br>(69,266 - 86,774) | 173,887<br>(156,162 - 192,231) |
| Scenario 5 | 74,919<br>(66,529 - 83,386) | 79,926<br>(70,776 - 89,192) | 154,845<br>(138,789 - 171,916) |
| Scenario 6 | 149,649<br>(133,203 - 166,751) | 160,061<br>(142,105 - 178,186) | 309,711<br>(276,892 - 342,894) |

*Scenario 1: Reducing current salt intake to an average of 5 g/day (Australian Suggested Dietary Target), Scenario 2: 30% relative reduction in current salt intake (National Preventive Health Strategy, 2021-2030 target), Scenario 3: 20% relative reduction in current salt intake, Scenario 4: 15% relative reduction in current salt intake; Scenario 5: reduction in current salt intake by 1g; Scenario 6: reduction in current salt intake by 2g. UI, uncertainty intervals.*

**Figure 1:**
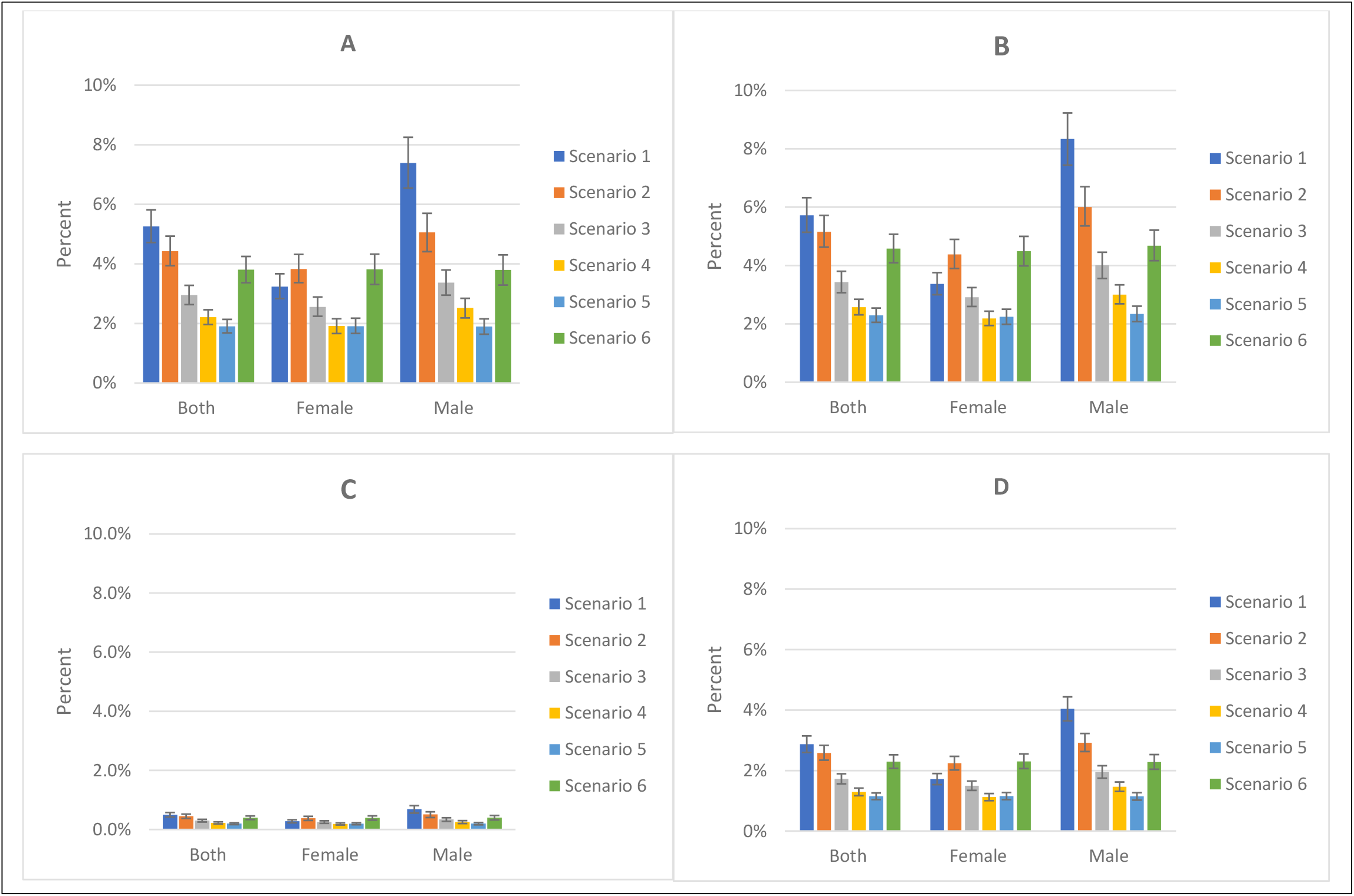
Relative reductions in incidence (panel A, 2019 – 2030; panel B, lifetime) and mortality (panel C, 2019 – 2030; panel D, lifetime)

### Estimated changes in CKD mortality by 2030 and lifetime

Over 11 years, achieving the Australian SDT could prevent 570 (95%UI: 479 to 652) deaths from CKD while reaching the NPHS target by 2030 could prevent 511 (95%UI: 426 to 590) CKD deaths. Over the remaining lifetime, over 22,500 (20,400 to 24,750) and 20,290 (18,440 to 22,275) deaths for SDT and NPHS targets respectively could be averted. In the two scenarios and over both time horizons, CKD due to hypertension (40%) and CKD due to glomerulonephritis (30%) had the largest deaths averted. Estimated relative reductions in cumulative CKD mortality over the lifetime of women were 1.7% for the SDT and 2.2% for the NPHS, while over the lifetime of men, reductions were 4.0% for the SDT and 2.9% for the NPHS. See Table 3, Figure 1 and supplementary file Figures 2, 3, 6 and 7.

**Table 3:** Estimated reductions in cumulative number of deaths from CKD in Australia between 2019 to 2030, and over the lifetime.

| Reductions between 2019 to 2030 |  |  |  |
| --- | --- | --- | --- |
|  | Male<br>(Mean 95% UIs) | Female<br>(Mean 95% UIs) | Both<br>(Mean 95% UIs) |
| Scenario 1 | 423<br>(341 - 496) | 145<br>(118 - 171) | 568<br>(479 - 652) |
| Scenario 2 | 313<br>(249 - 368) | 198<br>(159 - 231) | 511<br>(426 - 590) |
| Scenario 3 | 209<br>(169 - 244) | 132<br>(107 - 153) | 341<br>(288 - 390) |
| Scenario 4 | 157<br>(124 - 185) | 99<br>(79 - 116) | 255<br>(214 - 293) |
| Scenario 5 | 124<br>(98 - 146) | 103<br>(84 - 120) | 226<br>(188 - 260) |
| Scenario 6 | 248<br>(199 - 292) | 205<br>(163 - 240) | 453<br>(381 - 520) |
| Reductions over the remaining lifetime |  |  |  |
|  | Male<br>(Mean 95% UIs) | Female<br>(Mean 95% UIs) | Both<br>(Mean 95% UIs) |
| Scenario 1 | 15,798<br>(14,248 - 17,362) | 6,785<br>(6,067 - 7,515) | 22,583<br>(20,414 - 24,752) |
| Scenario 2 | 11,425<br>(10,286 - 12,627) | 8,866<br>(7,975 - 9,768) | 20,291<br>(18,441 - 22,276) |
| Scenario 3 | 7,638<br>(6,836 - 8,455) | 5,913<br>(5,314 - 6,523) | 13,551<br>(12,262 - 14,891) |
| Scenario 4 | 5,730<br>(5,125 - 6,347) | 4,437<br>(3,965 - 4,902) | 10,168<br>(9,189 - 11,168) |
| Scenario 5 | 9,047<br>(8,176 - 9,913) | 4,565<br>(4,103 - 5,035) | 4,481<br>(3,995 - 4,963) |
| Scenario 6 | 8,924<br>(7,995 - 9,900) | 9,108<br>(8,162 - 10,077) | 18,032<br>(16,295 - 19,838) |

*Scenario 1: Reducing current salt intake to an average of 5 g/day (Australian Suggested Dietary Target), Scenario 2: 30% relative reduction in current salt intake (National Preventive Health Strategy, 2021-2030 target), Scenario 3: 20% relative reduction in current salt intake, Scenario 4: 15% relative reduction in current salt intake; Scenario 5: reduction in current salt intake by 1g; Scenario 6: reduction in current salt intake by 2g. UI, uncertainty intervals.*

### Life years and healthcare costs

Achieving the SDT and NPHS targets were estimated to result in 4,645 CKD-related HALYs and 3,870 CKD-related HALYs gained respectively by 2030, while over the remaining lifetime, we project 200,000 HALYs could be gained for the SDT and 170,000 HALYs for the NPHS target. For health expenditure, achieving the SDT and NPHS target could save AU$ 57 million and AU$ 44 million respectively in CKD-related health care costs by 2030. Over the lifetime, these savings increased substantially to AU$ 664 million and AU$ 514 million respectively. See Figure 2.

**Figure 2:**
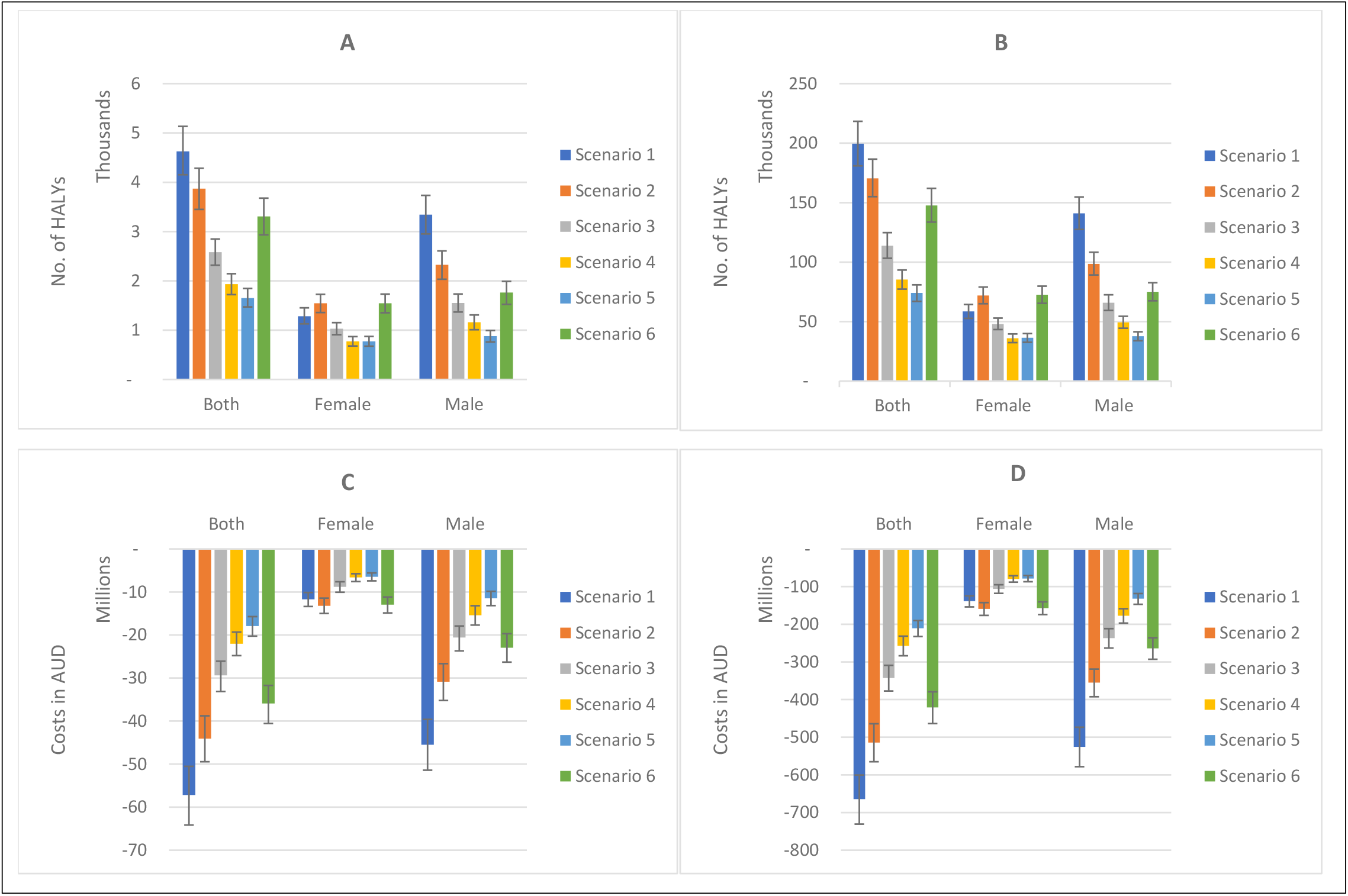
HALYs gained (panel A, 2019 – 2030; panel B, lifetime) and CKD health expenditure savings (panel C, 2019 – 2030; panel D, lifetime)

### Scenario analyses

In the main analysis, we present undiscounted HALYs, and costs were discounted at 3%. Applying a 3% discount rate, HALYs dropped by 22.5% in the first 11 years, and reduced by 67.6% over the lifetime. When a 5% discount rate was applied to costs, savings in health expenditure declined by 15.8% in the first 11 years and by 43% over the lifetime. We also modelled the impact of other modest reductions in sodium. For example, a 15% sodium reduction (i.e., half of the NPHS target, but similar to the United Kingdom salt reduction program effect) over 11 years, could prevent 24,900 new CKD cases and avert 255 CKD deaths. This translates to 85,320 CKD-related HALYs gained over their remaining lifetime. Furthermore, if only 1g of salt (∼390mg sodium) was reduced, that is, from 9.6g of salt (3,700mg of Na) to 8.6g of salt (3,310mg of Na), there would be 21,400 fewer new cases of CKD and 225 deaths averted by 2030. Over the lifetime, these increase by 7 times for incidence and 19 times for deaths and results in 74,000 CKD-related HALYs gained. See supplementary file Figures 8 – 10.

## DISCUSSION

This study projected the impact of achieving the National Preventive Health Strategy 2021-2030 and the Suggested Dietary Targets for sodium intake on the burden of CKD in Australia. Based on current salt and BP distributions for people aged 25 years and above, our modelling shows that achieving the NPHS sodium target could result in 49,800 fewer new cases of CKD and 509 deaths averted by 2030. Furthermore, attaining the Australian SDT for sodium could prevent over 59,000 new CKD cases and postpone 570 CKD deaths by 2030 in Australia. If the target sodium levels are sustained for the remaining lifetime, these impacts substantially increase (about 7 times for incidence and 40 times for deaths), translating to 165,000 HALYs (for NPHS) and 200,000 HALYs (for SDT) gained, with considerable savings in health expenditure. As expected, discounting HALYs and higher discount rate on costs had a significant impact on outcomes. These novel findings demonstrate the potential extra benefits of sodium reduction besides its impact on heart disease and stroke.

### Comparison with other studies

We estimate that achieving the NPHS and SDT for sodium intake could reduce average systolic BP of adult Australians by 1.7 mmHg and 2.1 mmHg respectively. High certainty evidence from a recent Cochrane meta-analysis showed that salt reduction of 4.2g/day (1,640mg of sodium) in people with CKD decreased systolic BP by 6.9 mmHg, and also reduced albuminuria by 36% (10). Studies have shown that the effect of high sodium intake on BP are greater in people with CKD compared to the general non-CKD population, due to decreased renal excretion of the excess sodium (8, 33). As a result, reducing sodium intake is a powerful tool to achieve blood pressure control in people with CKD thus decelerating disease progression. Moreover, impaired kidney function is itself a prognostic factor for poor CVD outcomes (8). Thus, population sodium reduction efforts that prevent CKD potentially deliver extra benefits via a ripple effect on CVD burden.

Our literature search identified only one study that has modelled the long-term impacts of sodium reduction on CKD burden (12). In this Dutch study, Hendriksen et al. used baseline data from the PREVEND study to develop a Markov simulation model. Their projections showed that achieving a 6g/day target of salt led to 290,000 fewer cases of CKD (1.1% reduction) and 470 fewer cases of ESKD (3.2% reduction) after 20 years. A recent modelling study in Australia found that full compliance with the Australia’s sodium reformulation program could prevent 565 new CKD cases, avert 40 CKD deaths and 527 CKD-related DALYs a year (20). These findings are lower than our study results and is likely due to the following reasons. First, the magnitude of sodium reduction achieved from the reformulation program was 107 mg/day, which though similar to the annual reduction for the NPHS target (110mg/day), is lower than the annual sodium reduction from the SDT (180mg/day) modelled in our study. Second, their study used comparative risk assessment modelling to estimate the disease burden attributable to sodium. This framework lacks the time component and thus estimates were for a single year. In contrast, our dynamic multistate lifetable Markov model allowed for more nuanced time-dependent projections with linear incremental annual reductions over 11 years (through to 2030) and impacts over the lifecourse. The differences in modelling frameworks, differences in intervention effects and time horizon likely explain the difference in health outcomes. However, these studies demonstrate the additional benefits of sodium reduction besides its effect on CVDs.

### Implications of the findings

The Commonwealth Government of Australia has endorsed the WHO global sodium reduction recommendation. However, recent evidence suggests that Australia is not on track to achieve this 30% reduction target by 2025 (34). Our modelled projections demonstrate the potential CKD-related population health gains that could be missed if current dietary salt intake patterns persist. Moreover, in addition to the avoidable morbidity that could mitigate increasing hospitalizations and demand for renal replacement therapy (35), significant savings in health expenditure could be obtained. These funds could be invested to further strengthen preventive health efforts or other social causes to improve the wellbeing of Australians.

Most of the sodium in the Australian food system is from pre-packaged and ultra-processed foods (36), hence food reformulation is a viable option to reduce salt intake in the population. The Australian Government Healthy Food Partnership has rightfully set sodium reformulation targets for some of these foods (37); however, these targets are far from comprehensive. Recent modelling indicates that much larger (more than double) health gains could be achieved if the range of products were as extensive as that implemented in the United Kingdom reformulation program (20). To maximise impact, there is an urgent need to expand the range of products in this program perhaps through adapting the broad benchmarks recently set by the WHO (38) and making reformulation mandatory. Additionally, there should be regular revision to lower sodium targets in a framework to achieve the National Preventive Health Strategy target of a 30% reduction in sodium intake by 2030, and further down to the Australian SDT average salt intake of 5g/day (2,000mg sodium) (21, 22).

Our study shows that achieving the SDT (akin to the WHO recommendation) could deliver greater health benefits. Government inaction or delays in enforcing strong policies can have substantial health and economic costs. A recent analysis of the United States Food and Drug Administration’s 4.3-year delay (2016 to 2021) in finalizing their short-term sodium reduction targets was estimated to cost over 250,000 lives over 10 years (39). Federal and State Governments in Australia must therefore work together to enforce robust and timely policies to avoid such losses. Realizing the policy targets modelled here has the potential to achieve even bigger health gains, as other health outcomes related to excess sodium intake like heart disease, stroke and stomach cancer (40-42) that could be averted were beyond the scope of this work.

### Strengths and limitations

This study had some limitations. First, our projections do not incorporate trends in dietary sodium intake. Temporal changes in exposure are likely to influence our current projections. However, recent meta-regression of salt consumption in Australia did not find significant evidence of a change in sodium intake in the last 2 decades. As such, our assumption of stable intake seems conservative. Secondly, evidence suggests a direct impact of sodium on kidney function independent of BP (43). However, we modelled the effect mediated through BP, for which the evidence is strongest (7). It is possible that we may have underestimated the impact by not accounting for additional mechanisms by which sodium affects the kidneys. Third, impaired kidney function is a potent prognostic factor for poor CVD outcomes. While our study focused on CKD outcomes only, future modelling studies on the long-term impacts of sodium reduction and broader health outcomes should consider accounting for this additional effect. While this might not substantially change the overall recommendations for sodium reduction as a best-buy strategy to curb non-communicable diseases, accurate characterization of these pathophysiological mechanisms could refine modelled projections. Finally, our modelling does not account for socio-economic differences. Prior studies have suggested that preventive health strategies could have an impact on health inequalities (42), hence future work should consider evaluating the impacts of sodium reduction on health inequalities in Australia.

Our study has the following strengths. First, we maximise the use of Australia-specific data. Sodium intake was based on meta-analysis of surveys conducted across Australia including the 2011-12 National Nutrition and Physical Activity Survey, corrected for potential underestimation in dietary surveys to obtain weighted 24-hour urine estimates. We also used measured BP data from the Australian National Health Survey 2017-18. Use of these nationally representative data sources enhances the contextual relevance of our study. Secondly, our dynamic proportional multi-state lifetable model explicitly models age- and sex-specific cohorts of the population thus accounting for the varying impacts in sodium intake and BP distributions. In addition, we account for the differential impact of salt reduction in people with and without hypertension in line with evidence of the stronger effects of sodium on BP in those with hypertension (4, 7, 26). Third, there is limited empirical evidence on the longterm effects of sodium intake on CKD, echoed in a recent Cochrane meta-analysis (10). Our modelling compliments this evidence gap by projecting beyond intermediate outcomes (blood pressure) to CKD for the first time in Australia.

## CONCLUSION

This modelling study suggests that attaining the Australian National Preventive Health Strategy 2021-2030 and the NHMRC Suggested Dietary Targets for sodium intake could prevent a substantial CKD burden in Australia and lead to savings in health expenditure. Federal and State Governments need to step-up efforts to reduce sodium in the Australian food supply to curb related health and economic losses in the future. Further studies are warranted to evaluate the impacts of these policy targets on health inequality.

## Data Availability

All data used in this manuscript are publicly available and can be obtained from: Global Burden of Disease Results tool (https://ghdx.healthdata.org/gbd-results-tool), and from the Australian Bureau of Statistics National Health Survey 2017-18 (https://www.abs.gov.au/statistics/health/health-conditions-and-risks/national-health-survey-first-results/latest-release) and National Nutrition and Physical Activity Survey 2011-12 (https://www.abs.gov.au/statistics/microdata-tablebuilder/available-microdata-tablebuilder/australian-health-survey-nutrition-and-physical-activity).

