## Supplementary file for "Projected impact on blood pressure, chronic kidney disease burden and healthcare costs of achieving the Australian sodium reduction targets: a modelling study"

A. **Input data**
1. **Salt intake (grams/day)**

|  | **Men** | | **Women** | |
| --- | --- | --- | --- | --- |
| **Age groups** | **Mean** | **95% CI** | **Mean** | **95% CI** |
| 19-30 years | 11.68 | 11.23 – 12.13 | 8.24 | 7.84 – 8.65 |
| 31-50 years | 10.91 | 10.49 – 11.33 | 7.71 | 7.33 – 8.09 |
| 51-70 years | 9.39 | 9.03 – 9.76 | 7.06 | 6.71 – 7.41 |
| 71+ years | 8.30 | 7.98 – 8.62 | 6.35 | 6.04 – 6.66 |
| Overall | 10.07 | 9.68 – 10.46 | 7.34 | 6.98 – 7.70 |

Estimates are weighted mean salt intake obtained from 24-hour urine collections after adjusting for non-urinary salt excretion. These are from a meta-analysis [1] that included 31 published and 1 unpublished dataset of studies conducted across Australia between 1989 and 2015. This meta-analysis included the National Nutrition and Physical Activity Survey (NNPAS) 2011-12 [2] from which we derived the age pattern of sodium consumption and used to adjust the overall estimates accordingly. *CI, confidence interval.*

2. **Systolic blood pressure (mmHg)**

|  | **Men** | | **Women** | |
| --- | --- | --- | --- | --- |
| **Age group** | **Mean (SD)** | **RSE, %** | **Mean (SD)** | **RSE, %** |
| 18 – 24 years | 119.5 (18.2) | 0.7 | 107.6 (14.7) | 0.5 |
| 25 – 34 years | 120.3 (13.5) | 0.4 | 108.5 (15.7) | 0.4 |
| 35 – 44 years | 121.3 (11.6) | 0.4 | 112.5 (15.7) | 0.4 |
| 45 – 54 years | 126.5 (17.9) | 0.4 | 119.6 (17.0) | 0.5 |
| 55 – 64 years | 132.4 (14.7) | 0.5 | 126.8 (20.8) | 0.5 |
| 65 – 74 years | 134.9 (21.3) | 0.5 | 133.6 (17.7) | 0.5 |
| 75 – 84 years | 136.5 (20.9) | 0.7 | 137.9 (17.0) | 0.6 |
| 85+ years | 140.2 (33.3) | 1.3 | 140.8 (28.0) | 1.2 |
| Overall | 126.1 (16.8) | 0.2 | 119.2 (21.0) | 0.2 |

Data are based on measured blood pressure from the National Health Survey, 2017-18: First Results conducted by the Australian Bureau of Statistics [3]. This survey reported mean blood pressures, the proportion of people with hypertension and the proportion of people in various blood pressure ranges, e.g., <100mmHg, 100 to ≤110 mmHg, etc. together with the relative standard error (RSE) of the estimates. We used the RSE to back-calculate the standard errors, and assuming a normal distribution, we used the proportions of people in various blood pressure categories to derive the variance and hence standard deviations (SD).

3. **Relative risks**

| **Age** | **CKD-HTN** | **CKD-DM** | **CKD-GMN** | **CKD-U** |
| --- | --- | --- | --- | --- |
| 25-29yrs | 1.281  (1.180 – 1.385) | 1.283  (1.186 – 1.397) | 1.281  (1.182 to 1.383) | 1.282  (1.181 to 1.395) |
| 30-34yrs | 1.281  (1.180 – 1.385) | 1.283  (1.186 – 1.397) | 1.281  (1.182 to 1.383) | 1.282  (1.181 to 1.395) |
| 35-39yrs | 1.281  (1.180 – 1.385) | 1.283  (1.186 – 1.397) | 1.281  (1.182 to 1.383) | 1.282  (1.181 to 1.395) |
| 40-44yrs | 1.281  (1.180 – 1.385) | 1.283  (1.186 – 1.397) | 1.281  (1.182 to 1.383) | 1.282  (1.181 to 1.395) |
| 45-49yrs | 1.281  (1.180 – 1.385) | 1.283  (1.186 – 1.397) | 1.281  (1.182 to 1.383) | 1.282  (1.181 to 1.395) |
| 50-54yrs | 1.281  (1.180 – 1.385) | 1.283  (1.186 – 1.397) | 1.281  (1.182 to 1.383) | 1.282  (1.181 to 1.395) |
| 55-59yrs | 1.281  (1.180 – 1.385) | 1.283  (1.186 – 1.397) | 1.281  (1.182 to 1.383) | 1.282  (1.181 to 1.395) |
| 60-64yrs | 1.281  (1.180 – 1.385) | 1.283  (1.186 – 1.397) | 1.281  (1.182 to 1.383) | 1.282  (1.181 to 1.395) |
| 65-69yrs | 1.281  (1.180 – 1.385) | 1.283  (1.186 – 1.397) | 1.281  (1.182 to 1.383) | 1.282  (1.181 to 1.395) |
| 70-74yrs | 1.281  (1.180 – 1.385) | 1.283  (1.186 – 1.397) | 1.281  (1.182 to 1.383) | 1.282  (1.181 to 1.395) |
| 75-79yrs | 1.281  (1.180 – 1.385) | 1.283  (1.186 – 1.397) | 1.281  (1.182 to 1.383) | 1.282  (1.181 to 1.395) |
| 80-84yrs | 1.281  (1.180 – 1.385) | 1.283  (1.186 – 1.397) | 1.281  (1.182 to 1.383) | 1.282  (1.181 to 1.395) |
| 85-89yrs | 1.281  (1.180 – 1.385) | 1.283  (1.186 – 1.397) | 1.281  (1.182 to 1.383) | 1.282  (1.181 to 1.395) |
| 90-94yrs | 1.281  (1.180 – 1.385) | 1.283  (1.186 – 1.397) | 1.281  (1.182 to 1.383) | 1.282  (1.181 to 1.395) |
| 95+yrs | 1.281  (1.180 – 1.385) | 1.283  (1.186 – 1.397) | 1.281  (1.182 to 1.383) | 1.282  (1.181 to 1.395) |

Estimates are relative risks (and 95% uncertainty intervals) of developing chronic kidney disease (CKD) are from the Global Burden of Disease 2019 study [4]. These are reported to represent the risk of morbidity/mortality of either form of CKD for both men and women for every 10mmHg increase in systolic blood pressure. We used these relative risks to model the risk of incidence of the different forms of CKD due to increased systolic blood pressure. CKD-HTN, chronic kidney disease due to hypertension; CKD-DM, chronic kidney disease due to diabetes mellitus; CKD-GMN, chronic kidney disease due to glomerulonephritis; CKD-U, chronic kidney disease due to other or unspecified causes.

4. **Baseline CKD epidemiological data**
   4.1. **Incidence rates**

Numbers are rates per 1 and are Australian estimates from the Global Burden of Disease 2019 study [4]. Reported here are outputs after processing through the DISMOD-II epidemiological software [5]. It applies causal differential equations to estimate case fatality rates that were unavailable, while preserving consistency in the overall disease epidemiology.

4.1.1. **Men**

| **Age** | **CKD-HTN** | **CKD-DM** | **CKD-GMN** | **CKD-U** |
| --- | --- | --- | --- | --- |
| 25-29yrs | 0.0000 | 0.0002 | 0.0000 | 0.0013 |
| 30-34yrs | 0.0000 | 0.0005 | 0.0000 | 0.0013 |
| 35-39yrs | 0.0001 | 0.0004 | 0.0001 | 0.0013 |
| 40-44yrs | 0.0001 | 0.0004 | 0.0001 | 0.0010 |
| 45-49yrs | 0.0001 | 0.0005 | 0.0001 | 0.0010 |
| 50-54yrs | 0.0002 | 0.0007 | 0.0001 | 0.0018 |
| 55-59yrs | 0.0003 | 0.0011 | 0.0001 | 0.0026 |
| 60-64yrs | 0.0006 | 0.0017 | 0.0001 | 0.0048 |
| 65-69yrs | 0.0016 | 0.0030 | 0.0002 | 0.0114 |
| 70-74yrs | 0.0034 | 0.0052 | 0.0003 | 0.0239 |
| 75-79yrs | 0.0059 | 0.0055 | 0.0003 | 0.0398 |
| 80-84yrs | 0.0076 | 0.0030 | 0.0005 | 0.0489 |
| 85-89yrs | 0.0074 | 0.0005 | 0.0010 | 0.0410 |
| 90-94yrs | 0.0061 | 0.0000 | 0.0016 | 0.0167 |
| 95+yrs | 0.0068 | 0.0000 | 0.0026 | 0.0063 |

4.1.2. **Women**

| **Age** | **CKD-HTN** | **CKD-DM** | **CKD-GMN** | **CKD-U** |
| --- | --- | --- | --- | --- |
| 25-29yrs | 0.0000 | 0.0003 | 0.0000 | 0.0023 |
| 30-34yrs | 0.0000 | 0.0006 | 0.0000 | 0.0020 |
| 35-39yrs | 0.0001 | 0.0005 | 0.0000 | 0.0015 |
| 40-44yrs | 0.0001 | 0.0004 | 0.0000 | 0.0012 |
| 45-49yrs | 0.0001 | 0.0004 | 0.0000 | 0.0012 |
| 50-54yrs | 0.0002 | 0.0007 | 0.0000 | 0.0022 |
| 55-59yrs | 0.0004 | 0.0011 | 0.0000 | 0.0038 |
| 60-64yrs | 0.0007 | 0.0018 | 0.0000 | 0.0075 |
| 65-69yrs | 0.0017 | 0.0029 | 0.0001 | 0.0167 |
| 70-74yrs | 0.0032 | 0.0045 | 0.0001 | 0.0325 |
| 75-79yrs | 0.0047 | 0.0044 | 0.0002 | 0.0515 |
| 80-84yrs | 0.0054 | 0.0022 | 0.0002 | 0.0619 |
| 85-89yrs | 0.0049 | 0.0004 | 0.0005 | 0.0492 |
| 90-94yrs | 0.0041 | 0.0000 | 0.0010 | 0.0168 |
| 95+yrs | 0.0052 | 0.0000 | 0.0015 | 0.0060 |

CKD-HTN, Chronic kidney disease due to hypertension; CKD-DM, Chronic kidney disease due to diabetes mellitus; CKD-GMN, Chronic kidney disease due to glomerulonephritis; CKD-U, Chronic kidney disease due to other or unspecified causes.

4.2. **Prevalence rates**

Numbers are rates per 1 and are Australian estimates from the Global Burden of Disease 2019 study [4]. Reported here are outputs after processing through the DISMOD-II epidemiological software [5]. It applies causal differential equations to estimate case fatality rates that were unavailable, while preserving consistency in the overall disease epidemiology.

**4.2.1. Men**

| **Age** | **CKD-HTN** | **CKD-DM** | **CKD-GMN** | **CKD-U** |
| --- | --- | --- | --- | --- |
| 25-29yrs | 0.0004 | 0.0014 | 0.0018 | 0.0189 |
| 30-34yrs | 0.0005 | 0.0034 | 0.0018 | 0.0252 |
| 35-39yrs | 0.0008 | 0.0059 | 0.0020 | 0.0317 |
| 40-44yrs | 0.0011 | 0.0079 | 0.0022 | 0.0374 |
| 45-49yrs | 0.0016 | 0.0102 | 0.0024 | 0.0416 |
| 50-54yrs | 0.0024 | 0.0132 | 0.0026 | 0.0481 |
| 55-59yrs | 0.0036 | 0.0175 | 0.0029 | 0.0581 |
| 60-64yrs | 0.0058 | 0.0241 | 0.0031 | 0.0742 |
| 65-69yrs | 0.0108 | 0.0350 | 0.0034 | 0.1083 |
| 70-74yrs | 0.0225 | 0.0540 | 0.0040 | 0.1807 |
| 75-79yrs | 0.0444 | 0.0800 | 0.0045 | 0.3010 |
| 80-84yrs | 0.0752 | 0.0993 | 0.0044 | 0.4423 |
| 85-89yrs | 0.1060 | 0.1056 | 0.0037 | 0.5563 |
| 90-94yrs | 0.1264 | 0.1039 | 0.0025 | 0.6130 |
| 95+yrs | 0.1363 | 0.0997 | 0.0020 | 0.6262 |

4.2.2. **Women**

| **Age** | **CKD-HTN** | **CKD-DM** | **CKD-GMN** | **CKD-U** |
| --- | --- | --- | --- | --- |
| 25-29yrs | 0.0004 | 0.0019 | 0.0013 | 0.0271 |
| 30-34yrs | 0.0006 | 0.0042 | 0.0015 | 0.0376 |
| 35-39yrs | 0.0008 | 0.0070 | 0.0016 | 0.0455 |
| 40-44yrs | 0.0011 | 0.0092 | 0.0018 | 0.0515 |
| 45-49yrs | 0.0016 | 0.0113 | 0.0019 | 0.0563 |
| 50-54yrs | 0.0024 | 0.0139 | 0.0019 | 0.0641 |
| 55-59yrs | 0.0037 | 0.0181 | 0.0020 | 0.0774 |
| 60-64yrs | 0.0062 | 0.0250 | 0.0021 | 0.1015 |
| 65-69yrs | 0.0118 | 0.0360 | 0.0022 | 0.1518 |
| 70-74yrs | 0.0234 | 0.0533 | 0.0025 | 0.2478 |
| 75-79yrs | 0.0422 | 0.0749 | 0.0026 | 0.3897 |
| 80-84yrs | 0.0657 | 0.0897 | 0.0025 | 0.5431 |
| 85-89yrs | 0.0876 | 0.0942 | 0.0019 | 0.6565 |
| 90-94yrs | 0.1011 | 0.0927 | 0.0012 | 0.7066 |
| 95+yrs | 0.1074 | 0.0887 | 0.0009 | 0.7151 |

CKD-HTN, Chronic kidney disease due to hypertension; CKD-DM, Chronic kidney disease due to diabetes mellitus; CKD-GMN, Chronic kidney disease due to glomerulonephritis; CKD-U, Chronic kidney disease due to other or unspecified causes.

4.3. **Case fatality rates**

Numbers are rates per 1 and are based on Australian estimates from the Global Burden of Disease 2019 study [4]. Reported here are outputs after processing age- and sex-specific incidence, prevalence and mortality rates (remission assumed to be zero) for CKD through DISMOD-II software [5]. The software applies differential equations to estimate case fatality rates that were unavailable, while preserving consistency in overall epidemiology.

4.3.1. **Men**

| **Age** | **CKD-HTN** | **CKD-DM** | **CKD-GMN** | **CKD-U** |
| --- | --- | --- | --- | --- |
| 25-29yrs | 0.0001 | 0.0000 | 0.0006 | 0.0000 |
| 30-34yrs | 0.0002 | 0.0000 | 0.0009 | 0.0000 |
| 35-39yrs | 0.0002 | 0.0000 | 0.0016 | 0.0001 |
| 40-44yrs | 0.0004 | 0.0000 | 0.0026 | 0.0001 |
| 45-49yrs | 0.0006 | 0.0001 | 0.0039 | 0.0002 |
| 50-54yrs | 0.0008 | 0.0002 | 0.0057 | 0.0002 |
| 55-59yrs | 0.0012 | 0.0003 | 0.0088 | 0.0003 |
| 60-64yrs | 0.0018 | 0.0004 | 0.0141 | 0.0004 |
| 65-69yrs | 0.0023 | 0.0006 | 0.0218 | 0.0006 |
| 70-74yrs | 0.0030 | 0.0008 | 0.0346 | 0.0007 |
| 75-79yrs | 0.0042 | 0.0011 | 0.0609 | 0.0009 |
| 80-84yrs | 0.0067 | 0.0017 | 0.1229 | 0.0013 |
| 85-89yrs | 0.0123 | 0.0032 | 0.2952 | 0.0022 |
| 90-94yrs | 0.0233 | 0.0066 | 0.7195 | 0.0037 |
| 95+yrs | 0.0361 | 0.0125 | 1.3292 | 0.0057 |

4.3.2. **Women**

| **Age** | **CKD-HTN** | **CKD-DM** | **CKD-GMN** | **CKD-U** |
| --- | --- | --- | --- | --- |
| 25-29yrs | 0.0001 | 0.0000 | 0.0006 | 0.0000 |
| 30-34yrs | 0.0001 | 0.0000 | 0.0009 | 0.0000 |
| 35-39yrs | 0.0001 | 0.0000 | 0.0014 | 0.0001 |
| 40-44yrs | 0.0002 | 0.0000 | 0.0019 | 0.0001 |
| 45-49yrs | 0.0003 | 0.0001 | 0.0028 | 0.0001 |
| 50-54yrs | 0.0004 | 0.0001 | 0.0044 | 0.0002 |
| 55-59yrs | 0.0007 | 0.0002 | 0.0072 | 0.0002 |
| 60-64yrs | 0.0010 | 0.0003 | 0.0114 | 0.0004 |
| 65-69yrs | 0.0013 | 0.0004 | 0.0169 | 0.0004 |
| 70-74yrs | 0.0016 | 0.0005 | 0.0274 | 0.0005 |
| 75-79yrs | 0.0024 | 0.0008 | 0.0526 | 0.0007 |
| 80-84yrs | 0.0044 | 0.0013 | 0.1187 | 0.0010 |
| 85-89yrs | 0.0095 | 0.0027 | 0.3238 | 0.0017 |
| 90-94yrs | 0.0213 | 0.0065 | 0.8617 | 0.0033 |
| 95+yrs | 0.0365 | 0.0136 | 1.6273 | 0.0053 |

CKD-HTN, Chronic kidney disease due to hypertension; CKD-DM, Chronic kidney disease due to diabetes mellitus; CKD-GMN, Chronic kidney disease due to glomerulonephritis; CKD-U, Chronic kidney disease due to other or unspecified causes.

5. **Disability weights by CKD cause and overall population disability weights**
   5.1. **Men**

| **Age** | **CKD-HTN** | **CKD-DM** | **CKD-GMN** | **CKD-U** | **Overall pYLD rate** |
| --- | --- | --- | --- | --- | --- |
| 25-29yrs | 0.1267 | 0.0015 | 0.0644 | 0.0063 | 0.1081 |
| 30-34yrs | 0.1176 | 0.0022 | 0.0765 | 0.0058 | 0.1142 |
| 35-39yrs | 0.0998 | 0.0032 | 0.0873 | 0.0054 | 0.1207 |
| 40-44yrs | 0.0923 | 0.0054 | 0.0971 | 0.0056 | 0.1269 |
| 45-49yrs | 0.0799 | 0.0091 | 0.1096 | 0.0066 | 0.1329 |
| 50-54yrs | 0.0730 | 0.0125 | 0.1186 | 0.0071 | 0.1434 |
| 55-59yrs | 0.0661 | 0.0157 | 0.1273 | 0.0073 | 0.1603 |
| 60-64yrs | 0.0536 | 0.0173 | 0.1309 | 0.0072 | 0.1823 |
| 65-69yrs | 0.0415 | 0.0172 | 0.1372 | 0.0069 | 0.2115 |
| 70-74yrs | 0.0337 | 0.0167 | 0.1249 | 0.0063 | 0.2438 |
| 75-79yrs | 0.0298 | 0.0167 | 0.1110 | 0.0065 | 0.2745 |
| 80-84yrs | 0.0299 | 0.0186 | 0.1038 | 0.0082 | 0.3069 |
| 85-89yrs | 0.0290 | 0.0185 | 0.0904 | 0.0104 | 0.3385 |
| 90-94yrs | 0.0270 | 0.0168 | 0.0940 | 0.0116 | 0.3692 |
| 95+yrs | 0.0309 | 0.0176 | 0.0965 | 0.0158 | 0.4117 |

5.2. **Women**

| **Age** | **CKD-HTN** | **CKD-DM** | **CKD-GMN** | **CKD-U** | **Overall pYLD rate** |
| --- | --- | --- | --- | --- | --- |
| 25-29yrs | 0.1355 | 0.0018 | 0.0862 | 0.0070 | 0.1338 |
| 30-34yrs | 0.1252 | 0.0026 | 0.0961 | 0.0065 | 0.1391 |
| 35-39yrs | 0.1142 | 0.0041 | 0.0999 | 0.0066 | 0.1455 |
| 40-44yrs | 0.1009 | 0.0063 | 0.1143 | 0.0058 | 0.1532 |
| 45-49yrs | 0.0794 | 0.0099 | 0.1107 | 0.0063 | 0.1580 |
| 50-54yrs | 0.0618 | 0.0124 | 0.1199 | 0.0068 | 0.1642 |
| 55-59yrs | 0.0505 | 0.0150 | 0.1231 | 0.0068 | 0.1757 |
| 60-64yrs | 0.0375 | 0.0159 | 0.1209 | 0.0064 | 0.1921 |
| 65-69yrs | 0.0284 | 0.0163 | 0.1217 | 0.0058 | 0.2148 |
| 70-74yrs | 0.0253 | 0.0161 | 0.1230 | 0.0056 | 0.2422 |
| 75-79yrs | 0.0232 | 0.0163 | 0.1214 | 0.0062 | 0.2730 |
| 80-84yrs | 0.0238 | 0.0170 | 0.1209 | 0.0075 | 0.3123 |
| 85-89yrs | 0.0240 | 0.0170 | 0.1218 | 0.0092 | 0.3538 |
| 90-94yrs | 0.0221 | 0.0154 | 0.1105 | 0.0099 | 0.4013 |
| 95+yrs | 0.0243 | 0.0243 | 0.1102 | 0.0129 | 0.4574 |

6. **Population numbers, all-cause mortality rates and healthcare costs**

|  | **Men** | | | **Women** | | |
| --- | --- | --- | --- | --- | --- | --- |
| **Age group** | **Population numbers** | **Mortality rate** | **Cost per prevalent CKD** | **Population numbers** | **Mortality rate** | **Cost per prevalent CKD** |
| 25-29yrs | 957,746 | 0.0007 | AU$ 249 | 949,146 | 0.0003 | AU$ 168 |
| 30-34yrs | 933,587 | 0.0009 | AU$ 246 | 959,375 | 0.0004 | AU$ 166 |
| 35-39yrs | 885,449 | 0.0011 | AU$ 239 | 896,946 | 0.0006 | AU$ 200 |
| 40-44yrs | 793,623 | 0.0016 | AU$ 294 | 802,591 | 0.0009 | AU$ 220 |
| 45-49yrs | 825,686 | 0.0022 | AU$ 387 | 854,071 | 0.0013 | AU$ 267 |
| 50-54yrs | 750,782 | 0.0033 | AU$ 429 | 785,017 | 0.0020 | AU$ 280 |
| 55-59yrs | 757,941 | 0.0050 | AU$ 469 | 790,197 | 0.0030 | AU$ 251 |
| 60-64yrs | 677,332 | 0.0076 | AU$ 468 | 714,617 | 0.0046 | AU$ 232 |
| 65-69yrs | 596,458 | 0.0121 | AU$ 498 | 631,045 | 0.0072 | AU$ 243 |
| 70-74yrs | 518,927 | 0.0201 | AU$ 371 | 539,265 | 0.0122 | AU$ 181 |
| 75-79yrs | 351,089 | 0.0344 | AU$ 358 | 383,275 | 0.0220 | AU$ 172 |
| 80-84yrs | 227,926 | 0.0617 | AU$ 323 | 277,175 | 0.0425 | AU$ 156 |
| 85-89yrs | 129,208 | 0.1103 | AU$ 309 | 184,648 | 0.0830 | AU$ 150 |
| 90-94yrs | 53,978 | 0.1172 | AU$ 309 | 99,014 | 0.1524 | AU$ 150 |
| 95+ yrs | 13,876 | 0.2117 | AU$ 309 | 34,246 | 0.2209 | AU$ 150 |

Population data are from the Australian Bureau of Statistics estimated national resident population by age and sex as of 30 June 2019 [6]. Costs estimates were calculated using data from the Australian Institute of Health and Welfare (AIHW) health expenditure for Australia, 2018-2019 [7].

B. **Additional methods description**

***Estimating risk of CKD based on shifts in systolic blood pressure distribution***

To model the effect of the sodium reduction interventions on systolic blood pressure (SBP) and subsequent CKD, we used the potential impact fraction (PIF) – an epidemiological method that quantifies the proportional change in disease incidence (or mortality) that results from a change in exposure to a risk factor – to estimate changes in CKD incidence. We modelled blood pressure as a continuous variable (assuming a normal distribution) using the ‘distribution shift’ method [8]. This calculation includes integrating SBP distributions with the relative risk function for each 5-year age-group and sex. Given the difference in the effects of sodium on blood pressure in people with and without hypertension, we modified the PIF formula as seen below.

$PIF= \frac{\int_{a}^{b} RR\left( x \right)P\left( x \right)dx - \left( \int_{a1}^{b1} RR\left( x \right)P*\left( x \right)dx + \int_{a2}^{b2} RR\left( x \right)P*\left( x \right)dx \right)}{\int_{a}^{b} RR\left( x \right)P\left( x \right)dx}$

Where:

x = SBP exposure levels, RR(x) = relative risk function, P(x) = original SBP distribution, P* = SBP distribution without the intervention, a = start integration limit (90mmHg), b = end integration limit (220mmHg), a1 = start integration limit for normotensive people (90mmHg), b1 = end integration limit for normotensive people (139.9mmHg), a2 = start integration limit for hypertensive people (140mmHg), b2 = end integration limit for hypertensive people (220mmHg).

For all sex and five-year age groups, these calculations assumed a theoretical minimum risk exposure level (TMREL) of SBP of 115 mmHg, which is the lowest level considered for elevated vascular risk [9].

Post intervention incidence of CKD (I*) is: I* = I x (1-PIF).

Where:

I = current incidence of CKD in the Australian population by sex and five-year age groups.

I* = new incidence of CKD after the intervention.

These PIF calculations were done for each of the CKD causes (CKD due to hypertension, CKD due diabetes mellitus, CKD due to glomerulonephritis, CKD due to other or unspecified causes).

***Chronic kidney disease Markov models***

We developed Markov sub-models for each of the four main CKD causes, that is, CKD due to hypertension, CKD due to diabetes mellitus, CKD due to glomerulonephritis and CKD due to other or unspecified causes, as implemented in the GBD study [10]. Each disease Markov model has four distinct health states, that is, healthy (alive without CKD), diseased (alive with CKD), death from (any of the) CKDs and death due to other causes, with death being an absorbing state. The movement of proportions of the populations between these health states is governed by transition hazards or probabilities (incidence, case fatality, remission). In this chronic disease model, remission was assumed to be zero. In addition, for all four CKD causes modelled, the starting population in diseased state consisted of the proportion of prevalent cases in the population for each sex and five-year age-group, and the rest were allocated to the healthy state. For these models, a 1-year cycle length was used for state membership and the Markov models were linked to the life table. The figure below depicts the cardiovascular disease models.

Incidence

Remission

Mortality

(Other cause)

Mortality

(Other cause)

Case fatality

**Healthy**

(Alive without CKD)

**Dead**

(From other cause)

**Diseased**

(Alive with CKD)

**Dead**

(From CKD)

**Figure 1:** Conceptual model for chronic kidney disease (adapted from Barendregt et al. [5])

***The proportional multi-cohort multistate life table model***

A proportional multi-cohort multistate life table model was developed to estimate the difference in life years lived by the Australian population aged 25 years and above. We compared a population that continues to consume sodium at current levels with an identical ‘intervention’ population that achieves the sodium reduction targets. The multistate lifetable has the ability to deal with comorbidity with each of the diseases is assumed to be independent [11]. The population is stratified in multiple cohorts by sex and five-year age groups. Disease-specific morbidity and mortality rates from the Markov models with and without the sodium reduction influence the all-cause disability and background mortality rates (hence, dealing with competing mortality risks) in the lifetable. The calculated years lived by each of the cohorts is adjusted for sub-optimal health or disability (a numerical quantification of the degree of health loss due to a disease) using estimates derived from the GBD 2019 study [4].

For each of the CVDs modelled, this sub-optimal health adjustment is estimated by dividing the sex and age-specific years lived with disability (YLD) for that disease by the corresponding prevalence numbers. This gives an average weight of the proportion of health knocked out by the disease. This ‘disability’ weight is further adjusted for poor health due to background diseases not included in the model using all-cause prevalent years lived with disability (pYLD) per capita (after excluding the diseases modelled). This generates health-adjusted life years (HALY), where one HALY is equivalent to a year of life in perfect health. The difference in HALYs between the populations receiving the sodium reduction intervention and those not receiving is indicative of the health benefit attributed to the intervention. All the population cohorts are simulated for their remaining lifetime till they reach 100 years or die.

C. **Additional results**

A

|  | CKD_HTN | CKD_DM | CKD_GMN | CKD_Other |
| --- | --- | --- | --- | --- |
| Scen 1 | 7555.95 | 10865.5 | 1042.64 | 21054.1 |
| Scen 2 | 5459.58 | 7273.84 | 710.101 | 14304.1 |
| Scen 3 | 3646.55 | 4860.14 | 473.029 | 9516.75 |
| Scen 4 | 2731.71 | 3637.49 | 354.348 | 7122.87 |
| Scen 5 | 2124.45 | 2686.39 | 264.931 | 5324.87 |
| Scen 6 | 4247.42 | 5367.83 | 531.045 | 10691.5 |

B

|  | CKD_HTN | CKD_DM | CKD_GMN | CKD_Other |
| --- | --- | --- | --- | --- |
| Scen 1 | 3466.5 | 4862.93 | 280.625 | 10094.2 |
| Scen 2 | 4417.77 | 5502.88 | 338.004 | 11883.3 |
| Scen 3 | 2949.49 | 3685.47 | 225.303 | 7913.66 |
| Scen 4 | 2211.83 | 2758.79 | 169.099 | 5932.75 |
| Scen 5 | 2256.9 | 2705.07 | 169.518 | 5886.82 |
| Scen 6 | 4496.83 | 5401.81 | 338.363 | 11812.8 |

C

Number

|  | CKD_HTN | CKD_DM | CKD_GMN | CKD_Other |
| --- | --- | --- | --- | --- |
| Scen 1 | 136.246 | 28.3187 | 198.547 | 59.7015 |
| Scen 2 | 101.937 | 20.3744 | 147.441 | 43.6818 |
| Scen 3 | 67.9873 | 13.6312 | 98.4931 | 29.1112 |
| Scen 4 | 50.9171 | 10.2111 | 73.7214 | 21.8016 |
| Scen 5 | 40.4034 | 7.90284 | 58.2152 | 17.0444 |
| Scen 6 | 80.9091 | 15.7934 | 116.852 | 34.2241 |

D

Number

|  | CKD_HTN | CKD_DM | CKD_GMN | CKD_Other |
| --- | --- | --- | --- | --- |
| Scen 1 | 46.6938 | 9.20585 | 65.4043 | 23.386 |
| Scen 2 | 64.3551 | 11.6622 | 90.666 | 31.0107 |
| Scen 3 | 42.8118 | 7.80185 | 60.4362 | 20.6165 |
| Scen 4 | 32.1488 | 5.83979 | 45.3485 | 15.4639 |
| Scen 5 | 33.5158 | 5.9406 | 47.4049 | 15.9187 |
| Scen 6 | 66.7634 | 11.8466 | 94.4513 | 31.8664 |

**Figure 2: Reduction in cumulative number of new CKD cases (panel A, men; panel B, women) and deaths (panel C, men; panel D, women) by CKD cause between 2019 to 2030.**

A

|  | CKD_HTN | CKD_DM | CKD_GMN | CKD_Other |
| --- | --- | --- | --- | --- |
| Scen 1 | 60443.9 | 58269.4 | 7776.51 | 140186 |
| Scen 2 | 44591.3 | 40971.1 | 5624.2 | 100953 |
| Scen 3 | 29846.3 | 27471.1 | 3764.45 | 67081.4 |
| Scen 4 | 22411.2 | 20606.8 | 2826.96 | 50222.1 |
| Scen 5 | 17748.6 | 15763.1 | 2211.91 | 39195.4 |
| Scen 6 | 35271.8 | 31359.8 | 4400.15 | 78617.7 |

B

|  | CKD_HTN | CKD_DM | CKD_GMN | CKD_Other |
| --- | --- | --- | --- | --- |
| Scen 1 | 27663 | 25753.8 | 2383.04 | 64332.6 |
| Scen 2 | 36895.5 | 31458.1 | 3178.14 | 84654.2 |
| Scen 3 | 24640.1 | 21074.2 | 2123.49 | 56014.9 |
| Scen 4 | 18507.3 | 15794.2 | 1597.01 | 41921.5 |
| Scen 5 | 19174.7 | 15934.2 | 1649.07 | 43168.5 |
| Scen 6 | 38120.3 | 31671.5 | 3288.66 | 86980.9 |

C

|  | CKD_HTN | CKD_DM | CKD_GMN | CKD_Other |
| --- | --- | --- | --- | --- |
| Scen 1 | 6112.27 | 1780.33 | 5036.33 | 2869 |
| Scen 2 | 4497.8 | 1244.61 | 3641.33 | 2041.35 |
| Scen 3 | 3009.68 | 834.039 | 2438.19 | 1355.72 |
| Scen 4 | 2259.96 | 625.385 | 1830.19 | 1014.9 |
| Scen 5 | 1786.89 | 476.544 | 1432.23 | 785.657 |
| Scen 6 | 3550.98 | 948.703 | 2847.54 | 1576.93 |

D

|  | CKD_HTN | CKD_DM | CKD_GMN | CKD_Other |
| --- | --- | --- | --- | --- |
| Scen 1 | 2849.27 | 899.917 | 1665.08 | 1371.1 |
| Scen 2 | 3790.89 | 1092.73 | 2216.62 | 1765.73 |
| Scen 3 | 2531.3 | 731.834 | 1481.91 | 1168.28 |
| Scen 4 | 1901.23 | 548.316 | 1113.62 | 874.301 |
| Scen 5 | 1968.68 | 552.114 | 1149.97 | 894.604 |
| Scen 6 | 3913.75 | 1097.98 | 2292.89 | 1803.03 |

**Figure 3: Reduction in cumulative number of new CKD cases (panel A, men; panel B, women) and deaths (panel C, men; panel D, women) by CKD cause over the remaining lifetime.**

scenario 1

Percentage reduction

|  | CKD_HTN | CKD_DM | CKD_GMN | CKD_Other | CKD_Overall |
| --- | --- | --- | --- | --- | --- |
| Both | 5.17755% | 5.97026% | 5.94042% | 4.95892% | 5.25609% |
| Female | 3.23935% | 3.70781% | 3.25043% | 3.04549% | 3.23464% |
| Male | 7.13643% | 8.21312% | 7.64289% | 7.09647% | 7.3871% |

scenario 2

Percentage reduction

|  | CKD_HTN | CKD_DM | CKD_GMN | CKD_Other | CKD_Overall |
| --- | --- | --- | --- | --- | --- |
| Both | 4.63967% | 4.84989% | 4.70524% | 4.16916% | 4.42782% |
| Female | 4.1283% | 4.19583% | 3.9151% | 3.58532% | 3.8292% |
| Male | 5.15649% | 5.49829% | 5.20535% | 4.82138% | 5.05889% |

scenario 3

Percentage reduction

|  | CKD_HTN | CKD_DM | CKD_GMN | CKD_Other | CKD_Overall |
| --- | --- | --- | --- | --- | --- |
| Both | 3.09835% | 3.24382% | 3.13502% | 2.77501% | 2.95284% |
| Female | 2.75624% | 2.8101% | 2.60968% | 2.38764% | 2.55498% |
| Male | 3.44413% | 3.67379% | 3.46752% | 3.20777% | 3.37225% |

scenario 4

Percentage reduction

|  | CKD_HTN | CKD_DM | CKD_GMN | CKD_Other | CKD_Overall |
| --- | --- | --- | --- | --- | --- |
| Both | 2.32213% | 2.42796% | 2.34991% | 2.07853% | 2.21162% |
| Female | 2.06691% | 2.10352% | 1.95868% | 1.78998% | 1.91486% |
| Male | 2.58007% | 2.74959% | 2.59753% | 2.40087% | 2.52446% |

scenario 5

Percentage reduction

|  | CKD_HTN | CKD_DM | CKD_GMN | CKD_Other | CKD_Overall |
| --- | --- | --- | --- | --- | --- |
| Both | 2.05805% | 2.04655% | 1.95039% | 1.78497% | 1.901% |
| Female | 2.10903% | 2.06257% | 1.96354% | 1.77613% | 1.9055% |
| Male | 2.00653% | 2.03067% | 1.94208% | 1.79485% | 1.89625% |

scenario 6

Percentage reduction

|  | CKD_HTN | CKD_DM | CKD_GMN | CKD_Other | CKD_Overall |
| --- | --- | --- | --- | --- | --- |
| Both | 4.10742% | 4.08806% | 3.90306% | 3.58282% | 3.8064% |
| Female | 4.20219% | 4.11879% | 3.91927% | 3.56407% | 3.81328% |
| Male | 4.01164% | 4.05759% | 3.89283% | 3.60375% | 3.79914% |

**Figure 4: Relative reduction in incidence of CKD by cause and gender for all scenarios between 2019 to 2030**

scenario 1

Percentage reduction

|  | CKD_HTN | CKD_DM | CKD_GMN | CKD_Other | CKD_Overall |
| --- | --- | --- | --- | --- | --- |
| Both | 6.67124% | 7.70427% | 7.07795% | 4.86515% | 5.72217% |
| Female | 4.17758% | 4.77032% | 4.02995% | 2.797% | 3.37254% |
| Male | 9.17761% | 10.5798% | 9.21779% | 7.36168% | 8.33794% |

scenario 2

Percentage reduction

|  | CKD_HTN | CKD_DM | CKD_GMN | CKD_Other | CKD_Overall |
| --- | --- | --- | --- | --- | --- |
| Both | 6.16866% | 6.64166% | 6.13274% | 4.41437% | 5.15221% |
| Female | 5.57049% | 5.8273% | 5.37321% | 3.67953% | 4.38391% |
| Male | 6.77018% | 7.44012% | 6.66675% | 5.30154% | 6.00768% |

scenario 3

Percentage reduction

|  | CKD_HTN | CKD_DM | CKD_GMN | CKD_Other | CKD_Overall |
| --- | --- | --- | --- | --- | --- |
| Both | 4.12501% | 4.45184% | 4.10254% | 2.928% | 3.43216% |
| Female | 3.72039% | 3.90399% | 3.59035% | 2.43498% | 2.91523% |
| Male | 4.53191% | 4.98903% | 4.46268% | 3.52324% | 4.00775% |

scenario 4

Percentage reduction

|  | CKD_HTN | CKD_DM | CKD_GMN | CKD_Other | CKD_Overall |
| --- | --- | --- | --- | --- | --- |
| Both | 3.09794% | 3.33829% | 3.08259% | 2.19187% | 2.57239% |
| Female | 2.79449% | 2.92598% | 2.70025% | 1.82242% | 2.18455% |
| Male | 3.40312% | 3.74259% | 3.35145% | 2.63792% | 3.00425% |

scenario 5

Percentage reduction

|  | CKD_HTN | CKD_DM | CKD_GMN | CKD_Other | CKD_Overall |
| --- | --- | --- | --- | --- | --- |
| Both | 2.79517% | 2.90703% | 2.69039% | 1.95904% | 2.29055% |
| Female | 2.89508% | 2.95201% | 2.78813% | 1.87652% | 2.2436% |
| Male | 2.69487% | 2.86306% | 2.62221% | 2.05867% | 2.34284% |

scenario 6

Percentage reduction

|  | CKD_HTN | CKD_DM | CKD_GMN | CKD_Other | CKD_Overall |
| --- | --- | --- | --- | --- | --- |
| Both | 5.5554% | 5.78016% | 5.35712% | 3.93818% | 4.58082% |
| Female | 5.75506% | 5.86701% | 5.55968% | 3.78044% | 4.49251% |
| Male | 5.35495% | 5.69527% | 5.21578% | 4.12861% | 4.67917% |

**Figure 5: Lifetime relative reduction in incidence of CKD by cause and gender for all scenarios**

scenario 1

Percentage reduction

|  | CKD_HTN | CKD_DM | CKD_GMN | CKD_Other | CKD_Overall |
| --- | --- | --- | --- | --- | --- |
| Both | 0.424164% | 0.421229% | 2.30322% | 0.167753% | 0.501452% |
| Female | 0.267194% | 0.220504% | 1.44518% | 0.0915823% | 0.279446% |
| Male | 0.531166% | 0.598305% | 2.86331% | 0.24875% | 0.688706% |

scenario 2

Percentage reduction

|  | CKD_HTN | CKD_DM | CKD_GMN | CKD_Other | CKD_Overall |
| --- | --- | --- | --- | --- | --- |
| Both | 0.385558% | 0.359656% | 2.07771% | 0.150803% | 0.451637% |
| Female | 0.368249% | 0.279372% | 2.00334% | 0.121453% | 0.381811% |
| Male | 0.397404% | 0.430484% | 2.1263% | 0.182012% | 0.510543% |

scenario 3

Percentage reduction

|  | CKD_HTN | CKD_DM | CKD_GMN | CKD_Other | CKD_Overall |
| --- | --- | --- | --- | --- | --- |
| Both | 0.256894% | 0.240615% | 1.38681% | 0.1004% | 0.301212% |
| Female | 0.244976% | 0.186896% | 1.33539% | 0.0807452% | 0.254291% |
| Male | 0.265049% | 0.288008% | 1.4204% | 0.1213% | 0.340796% |

scenario 4

Percentage reduction

|  | CKD_HTN | CKD_DM | CKD_GMN | CKD_Other | CKD_Overall |
| --- | --- | --- | --- | --- | --- |
| Both | 0.192593% | 0.180193% | 1.039% | 0.0752388% | 0.22572% |
| Female | 0.183961% | 0.139895% | 1.00202% | 0.0605649% | 0.190817% |
| Male | 0.198501% | 0.215746% | 1.06316% | 0.0908426% | 0.255164% |

scenario 5

Percentage reduction

|  | CKD_HTN | CKD_DM | CKD_GMN | CKD_Other | CKD_Overall |
| --- | --- | --- | --- | --- | --- |
| Both | 0.171385% | 0.155419% | 0.921634% | 0.0665526% | 0.200001% |
| Female | 0.191782% | 0.142315% | 1.04745% | 0.0623485% | 0.198501% |
| Male | 0.157511% | 0.166982% | 0.839542% | 0.0710231% | 0.201272% |

scenario 6

Percentage reduction

|  | CKD_HTN | CKD_DM | CKD_GMN | CKD_Other | CKD_Overall |
| --- | --- | --- | --- | --- | --- |
| Both | 0.342385% | 0.310312% | 1.84381% | 0.133435% | 0.400013% |
| Female | 0.382029% | 0.2838% | 2.08698% | 0.124809% | 0.395781% |
| Male | 0.315421% | 0.333705% | 1.68515% | 0.142609% | 0.403596% |

**Figure 6: Relative reduction in mortality from CKD by cause and gender for all scenarios between 2019 to 2030**

scenario 1

Percentage reduction

|  | CKD_HTN | CKD_DM | CKD_GMN | CKD_Other | CKD_Overall |
| --- | --- | --- | --- | --- | --- |
| Both | 3.02002% | 3.32658% | 5.60673% | 1.47444% | 2.87205% |
| Female | 2.00664% | 2.13626% | 3.29159% | 0.859948% | 1.71724% |
| Male | 3.94959% | 4.62798% | 7.30926% | 2.23589% | 4.0382% |

scenario 2

Percentage reduction

|  | CKD_HTN | CKD_DM | CKD_GMN | CKD_Other | CKD_Overall |
| --- | --- | --- | --- | --- | --- |
| Both | 2.79182% | 2.90103% | 4.90112% | 1.32312% | 2.57983% |
| Female | 2.66861% | 2.59462% | 4.38062% | 1.107% | 2.243% |
| Male | 2.90554% | 3.23615% | 5.28471% | 1.59093% | 2.92015% |

scenario 3

Percentage reduction

|  | CKD_HTN | CKD_DM | CKD_GMN | CKD_Other | CKD_Overall |
| --- | --- | --- | --- | --- | --- |
| Both | 1.86646% | 1.94361% | 3.28002% | 0.877389% | 1.72302% |
| Female | 1.78202% | 1.73775% | 2.9288% | 0.732597% | 1.49611% |
| Male | 1.94439% | 2.16877% | 3.53889% | 1.05681% | 1.95229% |

scenario 4

Percentage reduction

|  | CKD_HTN | CKD_DM | CKD_GMN | CKD_Other | CKD_Overall |
| --- | --- | --- | --- | --- | --- |
| Both | 1.40173% | 1.45691% | 2.46323% | 0.656777% | 1.29291% |
| Female | 1.33849% | 1.30204% | 2.20098% | 0.548291% | 1.12274% |
| Male | 1.4601% | 1.62629% | 2.65654% | 0.791208% | 1.46486% |

scenario 5

Percentage reduction

|  | CKD_HTN | CKD_DM | CKD_GMN | CKD_Other | CKD_Overall |
| --- | --- | --- | --- | --- | --- |
| Both | 1.26479% | 1.27688% | 2.16067% | 0.583981% | 1.15019% |
| Female | 1.38581% | 1.31117% | 2.27268% | 0.560977% | 1.155% |
| Male | 1.1542% | 1.23941% | 2.07879% | 0.612461% | 1.14542% |

scenario 6

Percentage reduction

|  | CKD_HTN | CKD_DM | CKD_GMN | CKD_Other | CKD_Overall |
| --- | --- | --- | --- | --- | --- |
| Both | 2.51373% | 2.54033% | 4.30085% | 1.17439% | 2.2923% |
| Female | 2.75478% | 2.60731% | 4.53098% | 1.13029% | 2.30392% |
| Male | 2.29346% | 2.46714% | 4.13258% | 1.22897% | 2.28074% |

**Figure 7: Lifetime relative reduction in mortality from CKD by cause and gender for all scenarios**

A

HALYs

|  | Scenario 1 | Scenario 2 | Scenario 3 | Scenario 4 | Scenario 5 | Scenario 6 |
| --- | --- | --- | --- | --- | --- | --- |
| Both | 3605.9 | 3016.49 | 1997.97 | 1498.35 | 1289.97 | 2580.94 |
| Female | 998.389 | 1206 | 796.988 | 599.129 | 602.69 | 1206.19 |
| Male | 2607.51 | 1810.49 | 1200.98 | 899.222 | 687.276 | 1374.74 |

B

HALYs

|  | Scenario 1 | Scenario 2 | Scenario 3 | Scenario 4 | Scenario 5 | Scenario 6 |
| --- | --- | --- | --- | --- | --- | --- |
| Both | 64686.1 | 54954.5 | 36457.6 | 27326.7 | 23712.1 | 47487 |
| Female | 18569.7 | 22757.6 | 15081.8 | 11320.8 | 11458.2 | 22939.4 |
| Male | 46116.3 | 32196.9 | 21375.8 | 16005.9 | 12254 | 24547.6 |

**Figure 8: HALYs (discounted at 3%) gained between 2019 to 2030 (panel A) and over the remaining lifetime (panel B)**

Scenario 1

Costs in AUD

|  | CKD_HTN | CKD_DM | CKD_GMN | CKD_Other | CKD_Overall |
| --- | --- | --- | --- | --- | --- |
| Both | -8173474 | -14013921 | -953148 | -25666427 | -48806970 |
| Female | -1577537 | -2761548 | -115493 | -5479135 | -9933713 |
| Male | -6595937 | -11252373 | -837655 | -20187292 | -38873257 |

Scenario 2

Costs in AUD

|  | CKD_HTN | CKD_DM | CKD_GMN | CKD_Other | CKD_Overall |
| --- | --- | --- | --- | --- | --- |
| Both | -6628407 | -10473825 | -679665 | -19780805 | -37562702 |
| Female | -1942509 | -3020447 | -125821 | -6209117 | -11297895 |
| Male | -4685899 | -7453377 | -553844 | -13571688 | -26264807 |

Scenario 3

Costs in AUD

|  | CKD_HTN | CKD_DM | CKD_GMN | CKD_Other | CKD_Overall |
| --- | --- | --- | --- | --- | --- |
| Both | -4391085 | -6931509 | -450253 | -13087462 | -24860308 |
| Female | -1285987 | -1994658 | -83188.2 | -4092076 | -7455910 |
| Male | -3105097 | -4936851 | -367064 | -8995386 | -17404398 |

Scenario 4

Costs in AUD

|  | CKD_HTN | CKD_DM | CKD_GMN | CKD_Other | CKD_Overall |
| --- | --- | --- | --- | --- | --- |
| Both | -3297212 | -5206241 | -335952 | -9826902 | -18666307 |
| Female | -965726 | -1504931 | -62325.4 | -3077742 | -5610725 |
| Male | -2331486 | -3701309 | -273627 | -6749160 | -13055582 |

Scenario 5

Costs in AUD

|  | CKD_HTN | CKD_DM | CKD_GMN | CKD_Other | CKD_Overall |
| --- | --- | --- | --- | --- | --- |
| Both | -2785205 | -4177016 | -262228 | -8076376 | -15300825 |
| Female | -977860 | -1456753 | -60458 | -3021728 | -5516799 |
| Male | -1807345 | -2720263 | -201770 | -5054649 | -9784027 |

Scenario 6

Costs in AUD

|  | CKD_HTN | CKD_DM | CKD_GMN | CKD_Other | CKD_Overall |
| --- | --- | --- | --- | --- | --- |
| Both | -5566452 | -8400535 | -525509 | -16199346 | -30691842 |
| Female | -1956318 | -2918873 | -121240 | -6074182 | -11070613 |
| Male | -3610134 | -5481662 | -404270 | -10125163 | -19621229 |

**Figure 9: Healthcare costs savings (discounted at 5%) between 2019 to 2030 by CKD cause and gender**

Scenario 1

Costs in AUD

|  | CKD_HTN | CKD_DM | CKD_GMN | CKD_Other | CKD_Overall |
| --- | --- | --- | --- | --- | --- |
| Both | -60867460 | -103157991 | -5856621 | -208153330 | -378035402 |
| Female | -12282061 | -20347258 | -671141 | -44827314 | -78127774 |
| Male | -48585399 | -82810733 | -5185479 | -163326017 | -299907628 |

Scenario 2

Costs in AUD

|  | CKD_HTN | CKD_DM | CKD_GMN | CKD_Other | CKD_Overall |
| --- | --- | --- | --- | --- | --- |
| Both | -49562458 | -77459669 | -4126948 | -160409841 | -291558916 |
| Female | -15110050 | -22554112 | -733213 | -51022036 | -89419411 |
| Male | -34452408 | -54905557 | -3393734 | -109387805 | -202139505 |

Scenario 3

Costs in AUD

|  | CKD_HTN | CKD_DM | CKD_GMN | CKD_Other | CKD_Overall |
| --- | --- | --- | --- | --- | --- |
| Both | -32927237 | -51331814 | -2732216 | -106045589 | -193036856 |
| Female | -10031748 | -14940681 | -485197 | -33716827 | -59174454 |
| Male | -22895489 | -36391133 | -2247019 | -72328762 | -133862402 |

Scenario 4

Costs in AUD

|  | CKD_HTN | CKD_DM | CKD_GMN | CKD_Other | CKD_Overall |
| --- | --- | --- | --- | --- | --- |
| Both | -24687221 | -38566661 | -2041068 | -79372005 | -144666954 |
| Female | -7520412 | -11260109 | -363664 | -25278907 | -44423093 |
| Male | -17166809 | -27306552 | -1677403 | -54093097 | -100243861 |

Scenario 5

Costs in AUD

|  | CKD_HTN | CKD_DM | CKD_GMN | CKD_Other | CKD_Overall |
| --- | --- | --- | --- | --- | --- |
| Both | -20891175 | -31067380 | -1580147 | -65150920 | -118689623 |
| Female | -7626718 | -10981586 | -353424 | -24891400 | -43853128 |
| Male | -13264457 | -20085795 | -1226723 | -40259520 | -74836494 |

Scenario 6

Costs in AUD

|  | CKD_HTN | CKD_DM | CKD_GMN | CKD_Other | CKD_Overall |
| --- | --- | --- | --- | --- | --- |
| Both | -41747780 | -62372241 | -3168062 | -131085415 | -238373499 |
| Female | -15245481 | -21986557 | -709311 | -50091673 | -88033021 |
| Male | -26502299 | -40385685 | -2458751 | -80993743 | -150340478 |

**Figure 10: Healthcare costs savings (discounted at 5%) over the remaining lifetime by CKD cause and gender**
